# FROM HEALTHCARE ACCESSIBILITY TO NATIONAL OUTPUT: HOW TECHNOLOGY-DRIVEN MRI IMPROVEMENTS CORRELATE WITH THE GROSS DOMESTIC PRODUCT

**DOI:** 10.1101/2025.10.05.25337388

**Authors:** Hsuan-Te Lee, Luen-Dau Li, Nan-kuei Chen

**Affiliations:** University of Arizona

**Keywords:** Magnetic Resonance Imaging (MRI), MRI Innovation, Healthcare Accessibility, Cobb–Douglas Production Function, Economic Growth, Gross Domestic Product (GDP)

## Abstract

Magnetic resonance imaging (MRI) is among the most transformative medical technologies; yet its widespread adoption in routine clinical practice remains constrained by high economic and technical barriers.. Recent advances in MRI—such as AI-enabled image reconstruction for accelerated MRI scans, cost-effective manufacturing, and the emergence of portable low-field scanners— are poised to improve accessibility and enhance diagnostic capacity. These advances are expected to be linked to national economic indicators such as the gross domestic product (GDP). This paper reviews recent innovations in MRI and analyzes their relationship to both population health and GDP. The main objective is to estimate the macroeconomic correlation, specifically GDP growth, associated with improved MRI accessibility and diagnostic capacity at the national healthcare and economic output level. Using a health-augmented Cobb–Douglas production framework and integrating data from existing clinical MRI reports into a simplified model, we conservatively estimate that expanded MRI accessibility could generate short-run national output growth in the range of **0.009515%** to **0.011678%**. Although these short-run gains appear statistically modest, the long-term implications are far more significant. Clinical innovations not only improve healthcare outcomes but also create substantial social and economic returns when considered over extended time horizons. Our findings underscore the importance of sustained investment and research in advanced diagnostic technologies such as MRI, highlighting their dual role in promoting public health and driving economic growth.

## 1. Introduction

The nexus of healthcare technology and economic performance has gradually become a critical discussion in the 21st century, particularly at present, as aging has become a crucial issue to address among both developing and developed nations (Masa Higo & Khan, 2014). Magnetic resonance imaging (MRI) is among the most disruptive tools in the imaging field, yet it is a massive financial challenge at the same time. Ever since it was first introduced to the clinic in the 1980s, MRI has revealed an entirely new world of diagnostics in the field of neurology and cardiology (Seiler et al., 2021; Constantine et al., 2004), as well as orthopedics and oncology. However, the initial high price, the special environment required, and maintenance also make it difficult to have such machines all over many places (Jalloul et al., 2023; Bayati et al., 2015), particularly in developing countries and underserved regions.

New technological advancements in healthcare machines came as a solution for the current circumstance. Image reconstruction can reduce scan time without sacrificing quality (Vranic et al., 2018; Farid GharehMohammadi & Sebro, 2024), and this allows more patients to be done in less time with the help of AI. Simultaneously, low-field MRI technology has resulted in portable devices that require minimal infrastructure and can operate within resource-deprived environments (Wald et al., 2019; Sneag et al., 2023; Alenezi et al., 2024; Wu & Feng, 2024). The cost is being lowered, and the machines are becoming easier to operate due to manufacturing advances. A combination of all these heads-up technologies provides us with an actual opportunity to equalize access to MRI and improve the diagnostic strength in the entire population.

The positive aspect of these advances is much more than improved health. By diagnosing sooner and minimizing uncertainty, we will save the cost of complications (Alenezi et al., 2024), which may chew into our budgets. At the macroeconomic level, there are fewer sick days, and days are worked more by healthier citizens who contribute to a stronger workforce (Jaana Kuoppala et al., 2008; Amiri & S. Behnezhad, 2020). The correlation between GDP growth and health infrastructure is established, and studies have continued to reveal that there are positive relations between healthcare and economic growth (Romaniuk et al., 2020; Park & Nam, 2019; Tanaka et al., 2021; Weil, 2005; Sterck et al., 2018).

Although it goes without saying that more appropriate diagnostics must contribute to the economic boost, there is almost no tangible evidence as to how the availability of MRI plays a role, if any, in supporting national productivity. The majority of the research considers the overall health outcomes or does not distinguish between different types of health infrastructure, but does not single out MRI. Not only MRI, but most of the clinical reports ignore the significance of healthcare treatments’ economic impact.

The gap that this project addresses is an investigation of how the technological advancements in MRI can influence the GDP of a country. We model a health-enhanced Cobb-Douglas production function, in which enhanced MRI capacity is related to economic growth. We use data on clinical outcomes and combine it with macroeconomic modeling to approximate the extent of productivity (and economy) benefits associated with MRI innovation.

The findings of this research can tell a lot to policymakers and investors. Countries all over the world are confronted by aging and constrained budgets (Sheiner, 2021); hence, an understanding of the actual economic payoffs to diagnostic technology has never been more vital than it is now. With the technological world continually reducing the cost of the highest level of imaging, the decision makers require well-founded, data-driven models to determine where to invest to achieve the best health and economic benefits.

## 2. Literature Review

### 2.1 Technology Advancement in MRI

Recent technological advances in MRI have enhanced diagnostic precision, improved patient accessibility, and streamlined clinical workflow. The most significant developments include, but are not limited to, AI-driven image reconstruction, high-field and cost-effective low-field MRI systems, improvement in contrast agents, and portable/cryogen-free MRI units.

**Figure 1.**
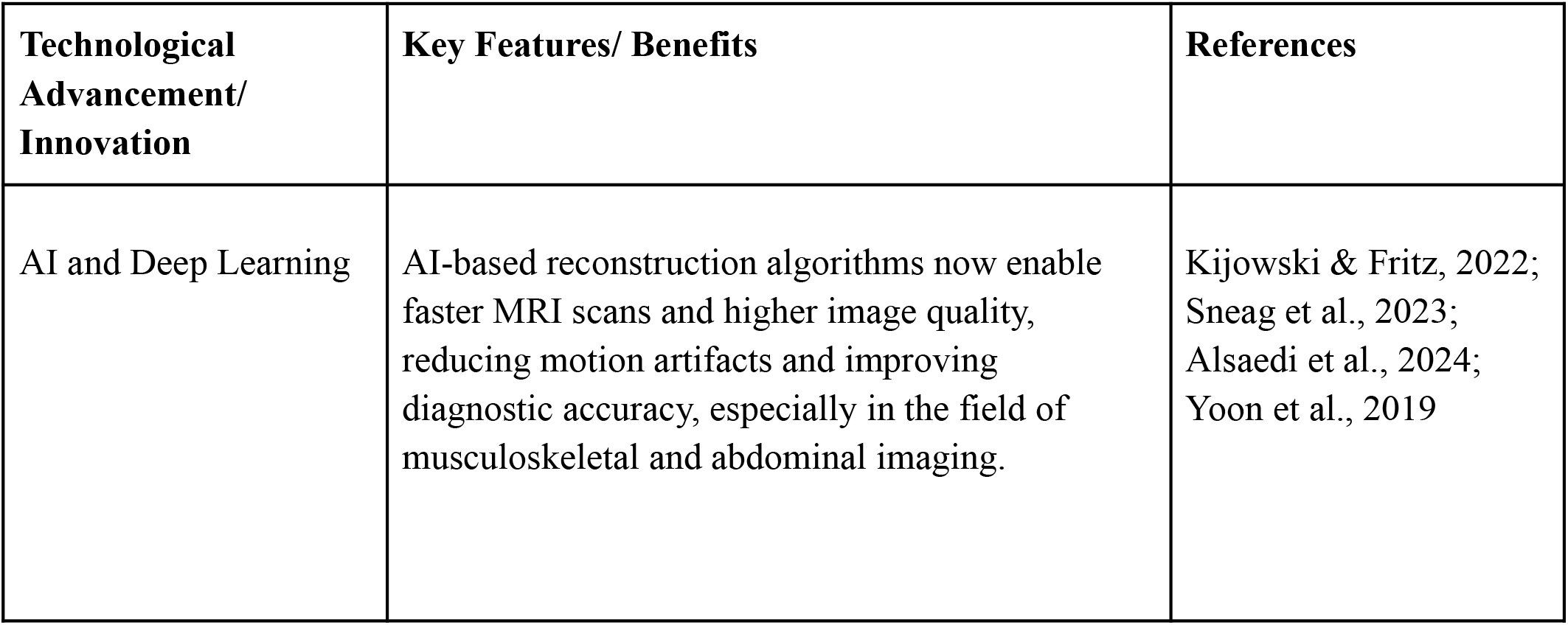
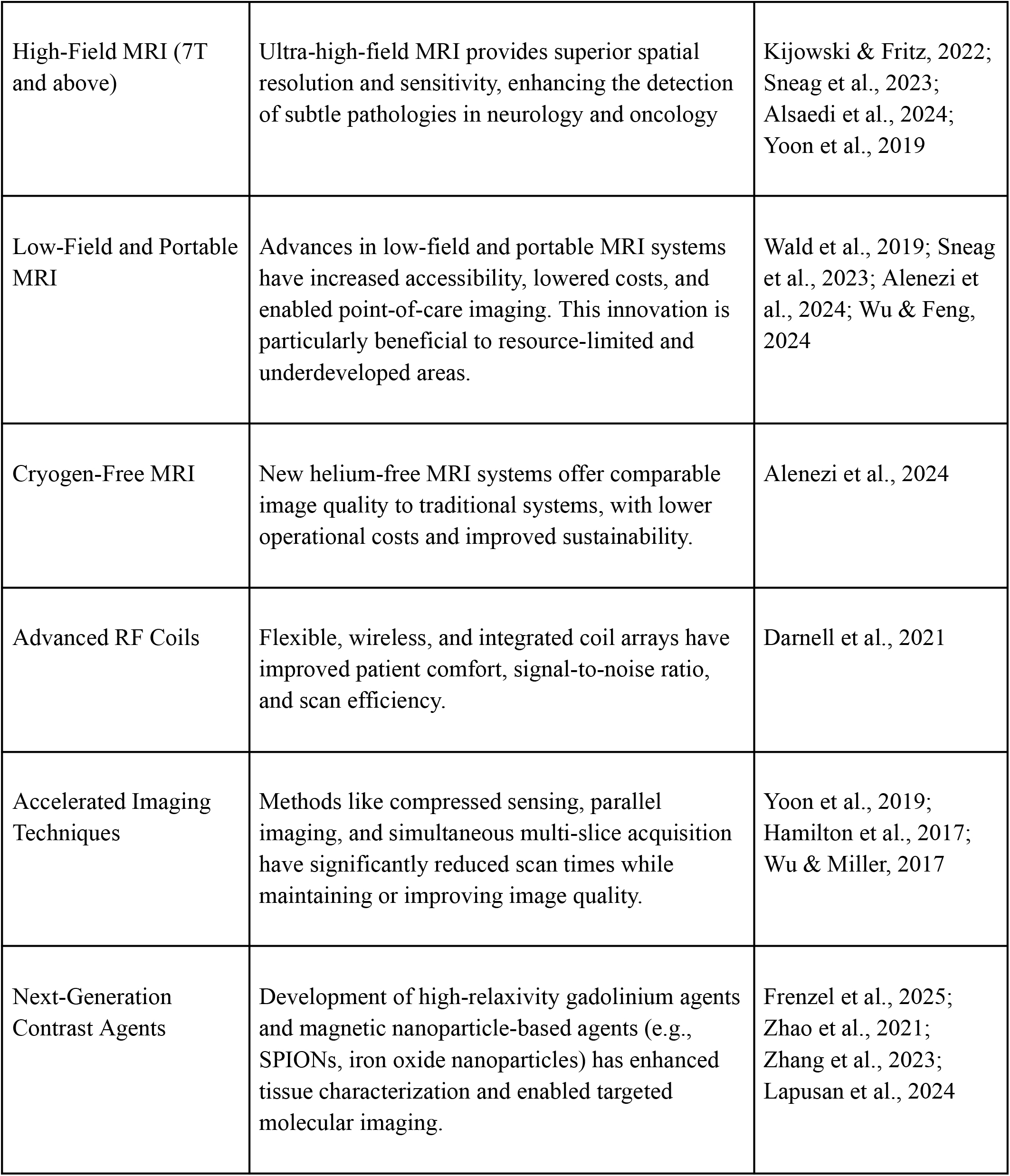
Major technology advancements and innovations in the past 10 years.

Summarizing the literature review above, it can be generalized that the qualitative importance of technological advancement in MRI has three broad effects on healthcare. To start with, new innovations in MRI have increased the accuracy of diagnosis and scanning. The higher resolution and faster imaging are useful in neurology, oncology, cardiovascular, as well as pediatric imaging (Alsaedi et al., 2024; Du et al., 2024; Sneag et al., 2023). Second, portable and affordable systems expand the use of MRI in underserved areas, gradually increasing the overall accessibility(Wald et al., 2019; Alenezi et al., 2024). Finally, more sophisticated imaging and targeted contrast agents are other tools that promote personalized diagnosis and treatment (Alsaedi et al., 2024; Zhang et al., 2023; Lapusan et al., 2024).

In conclusion, MRI technological innovations —especially in AI, hardware, and contrast agents— are revolutionizing clinical practice, offering better image quality, shorter scan times, and more access. Such innovations are driving MRI to individualized, effective, and available healthcare.

### 2.2 MRI Accessibility and Health Outcomes

In particular, with regard to the accessibility of MRI, greater MRI availability is linked to better national health, mostly by means of earlier diagnosis, less travel distance, and the facilitation of more equitable health care, but there are few solid quantitative data on the matter at the national level.

Based on multiple observational studies, countries with a higher MRI scanner density (scanners per million population) tend to have better health outcomes (more life expectancy and better disease management due to early and more precise diagnosis) (Geethanath & Vaughan, 2019; Hilabi et al., 2023). As an example, in Ghana, the access gap is 40 per cent with only 0.5 MRI scanners per million population, highlighting strong regional inequalities (Piersson & Gorleku, 2017). Optimization models used in Brazil suggest that 210 more MRI units would meet 95 percent of national demand, thus reducing the average patient travel to 44km in underserved regions, likely to improve access to care in a timely fashion and, accordingly, health outcomes (Almeida et al., 2022; De Freitas Almeida et al., 2022). In the United States, the distribution of MRI use decreases exponentially with poverty levels (*R*^2^ = 0.98), which indicates a close relationship between reduced access, increased poverty and poor health outcomes (Bermingham et al., 2024).

Although MRI is not a therapeutic intervention in itself, it has many effects on the overall health outcomes. Simply put, health outcomes can be traced to the augmentation of MRI accessibility in three main ways. First, more access to MRI will help identify and treat the disease earlier, eliminating needless treatments and hospitalization (Kang, 2019). Second, enhancing access to MRI in underserved areas reduces the impact of regional and socioeconomic health inequalities. Lastly, more efficient utilization of health care resources can be achieved through improved access because it will reduce the cost of treatment of diseases at later stages (Kang, 2019; De Freitas Almeida et al., 2022).

#### Quantitative Evidence Summary

**Figure 2.**
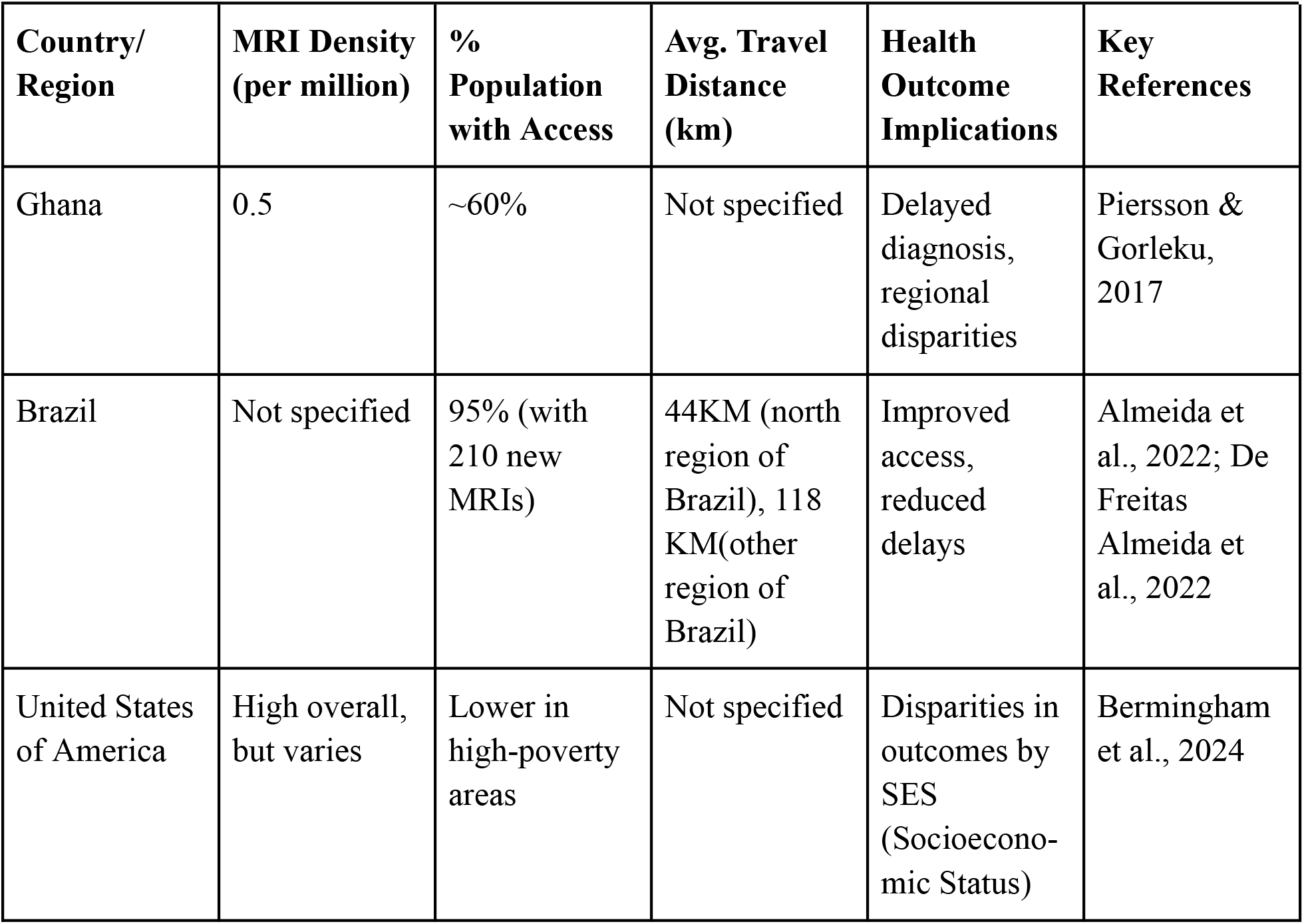
MRI access, travel distance, and health outcomes by country.

While direct national-level quantitative metrics are limited, multiple sources show that increased MRI accessibility improves health outcomes by enabling earlier diagnosis, reducing travel burdens, and promoting healthcare equity. Addressing disparities in MRI access is crucial for optimizing population health.

### 2.3 Health Outcomes and National Output

There is a positive correlation between health outcomes and national output; however, the degree of the association is dependent on a variety of economic, social, and governance factors. Numerous studies show that as the national output measured increases, the better the health indicators, such as the longer life expectancy and the lower infant and maternal mortality (Romaniuk et al., 2020; Bokhari et al., 2007; Park & Nam, 2019; Tanaka et al., 2021; Weil, 2005; Anwar et al., 2023; Rahman et al., 2018; Das, 2025; Onofrei et al., 2021; Sun et al., 2017) However, this association is still complicated and it is mediated by many other determinants.

**Figure 3.**
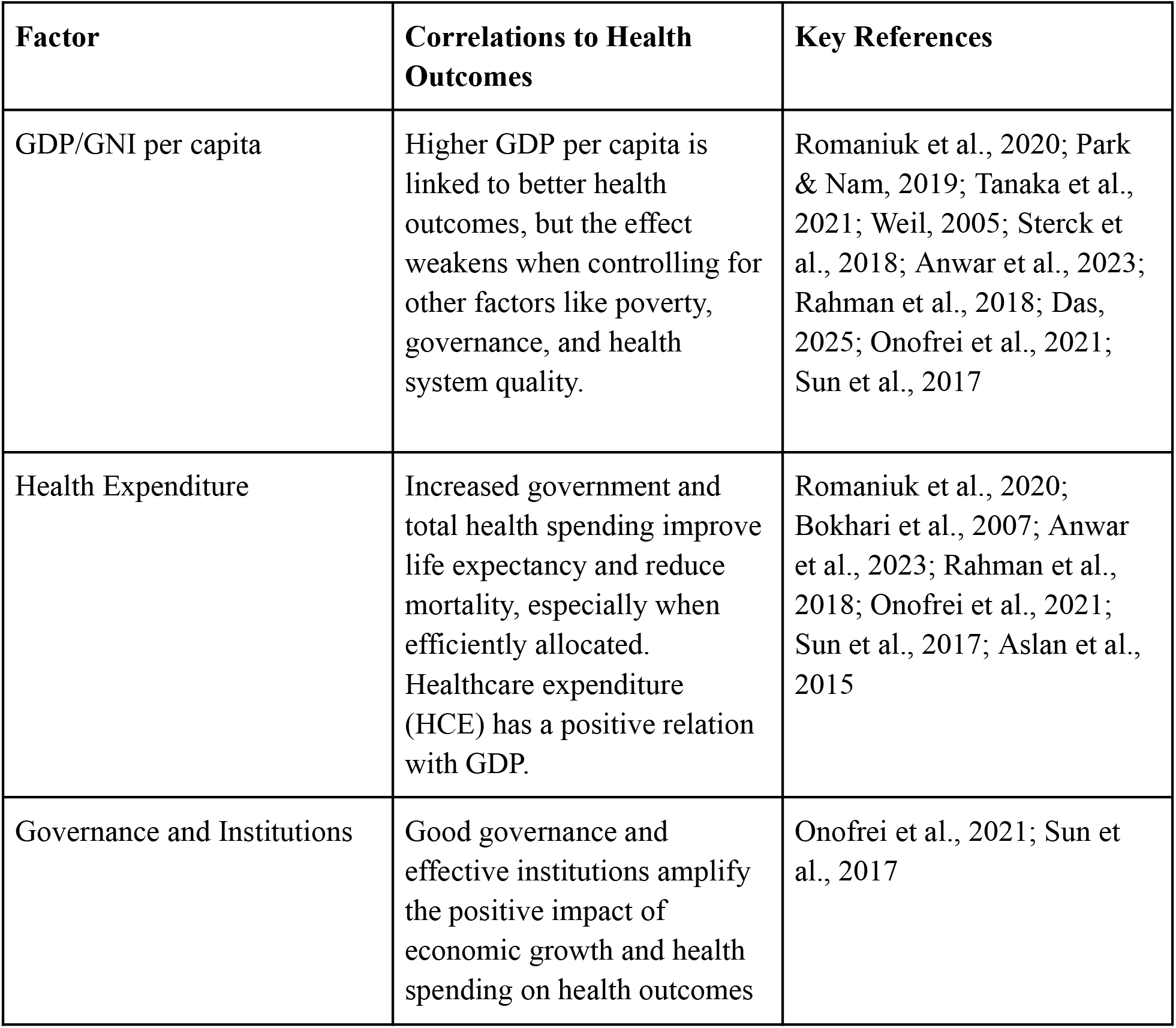
Table summarizing how national output and related factors affect health outcomes.

In general, the correlation between national output and health outcomes is strongest for basic indicators like infant mortality but weakens when accounting for poverty, governance, and epidemiological context (Sterck et al., 2018; Onofrei et al., 2021; Sun et al., 2017). In some regions, health improvements can drive economic growth in both developed countries and less developed nations (Kamanda et al., 2022; Weil, 2005; Aslan et al., 2015). Moreover, efficiency and allocation of health spending, not just the amount, are critical for translating economic gains into health improvements (Romaniuk et al., 2020; Anwar et al., 2023; Onofrei et al., 2021; Sun et al., 2017). Nonetheless, there are diminishing returns after a certain point; further increases in GDP have less impact on health outcomes (Spiteri & Von Brockdorff, 2019; Sterck et al., 2018).

As stated and demonstrated previously, health outcomes and national output are positively related. Economic growth can lead to a positive impact on health outcomes. Vice versa, enhancement in health outcome also has a positive impact on economic performance, despite being often overlooked.

In Weil’s Cobb–Douglas production function framework (Weil, 2005), health enters production through the labor composite *H*, where *H* = *h*·*v*·*L* (with *h* = human capital in the form of education, *v* = health-related human capital, and *L* = labor), so improvements in health raise the effective units of labor and thus increase aggregate output *Y* = *A·K*^*α*^·(*h·v·L*)^1−*α*^. In other words, better health (a higher *v* value) makes workers more productive—they work longer, exert more effort, and think more clearly—which raises the labor term; this increases output, generating additional indirect gains to GDP. Empirically, Weil shows that measurable health improvements (e.g., higher adult survival) translate into meaningful increases in labor input and hence GDP per worker.

## 3. Methodology

### 3.1 Theoretical Framework

The theoretical framework of the research is centred on the Cobb-Douglas production function. The concept of the Cobb-Douglas production function is to express economic output as the product of the quantitative factors of production along with their elasticity. In particular, the labor composite in the production function can be approximated through the product of the employment population times the percentage of workers who are in a healthy status. Extending this concept, we modeled a health-augmented Cobb-Douglas Aggragate production function to estimate the effect of improved accessibility in MRI (driven by technology advancement) on economic output: *Y* = *A·K*^*α*^·(*L·v*)^1−*α*^, where Y is the aggregate quantity of goods and services produced; A, conceptually, is the overall level of technology and efficiency in the production process; K is the value of physical assets used in production, such as machinery, buildings, and equipment; *v* stands for and interpret as the ratio of human labor in the form of health, where *v*∈[0, 1]; L is the labor composite, defined as the quantity of labor (the statistical employment population after adjusting mortality rate); *α* is the output elasticity of capital, which is how much output changes when capital input changes. (1 − *α*) is the theoretical value for the output elasticity of labor.

The approach of this research is to use the modeled health-augmented Cobb-Douglas Aggregate production function to approximate the impact of MRI accessibility (which improves the overall health level of a nation, v) on economic output. The independent variable will be the ratio of human labor in the form of health *v*, where *v*∈[0, 1], and the statistical employment population L. The dependent variable will be the relative percentage change in economic output, Y.

To simplify the statistical calculation, we consider the circumstance in the context of a hypothetical developing country that has no access to any MRI at first. The output of the hypothetical country before accessible to MRI is denoted as *Y*_*i*_(*A, K, L*_*i*_, *v*_*i*_)=*A·K*^α^ (*L*_*i*_·*vi*)^1−α^, and the output of the country after the ample accessibility of MRI is denoted as *Y*_*f*_(*A, K, L*_*f*_, *v*_*f*_)=*A·K*^α^ (*L*_*f*_·*v*_*f*_)^1−α^. Under the assumption that the relative change in *v* and L has a negligible impact on the other variables (A, K, and *α*), we calculate the relative percentage change in Y, which is real GDP in this case. 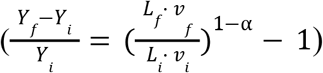

### 3.2 Data Sources and Variables

Based on the literature review on **2.2 MRI Accessibility and Health Outcomes**, an increase in accessibility to MRI does have a qualitatively positive impact on the variable v, defined as the ratio of human labor in the form of health (v∈[0,1]), yet the statistical evidence is insufficient throughout the research. Due to the research limitation, the variable v will be assumed to generally increase through the process of increasing MRI’s accessibility, where *v*_*f*_ *≥ v*_*i*_. Within the calculation process, we restrict the condition to *v*_*f*_ = *v*_*i*_.

The variable L, defined as the labour composite, will be estimated by the mortality improvement through pre- and post-implementation of MRI in the hypothetical country. L is defined such that *L* (*m*) = *E*·(1 − *m*), where E is the employed population before mortality adjustment (number of employed persons), m is the annual mortality rate of the employed population. In this research, we restrict attention to mortality data of breast cancer, traumatic brain injury (TBI), and ischemic stroke to produce a conservative, mortality-only lower-bound estimation of the effect. For the baseline, we (i) assume negligible comorbidity between these conditions (i.e., overlap in the same individual is negligible) (ii) focus on cases within the working population (we thus treat cases outside the labour force as negligible) (iii) a sex ratio of 1:1 for the employment population. (iv) the employment population before mortality adjustment *E* remains constant through the time interval of MRI implementation.

**Figure 4.**
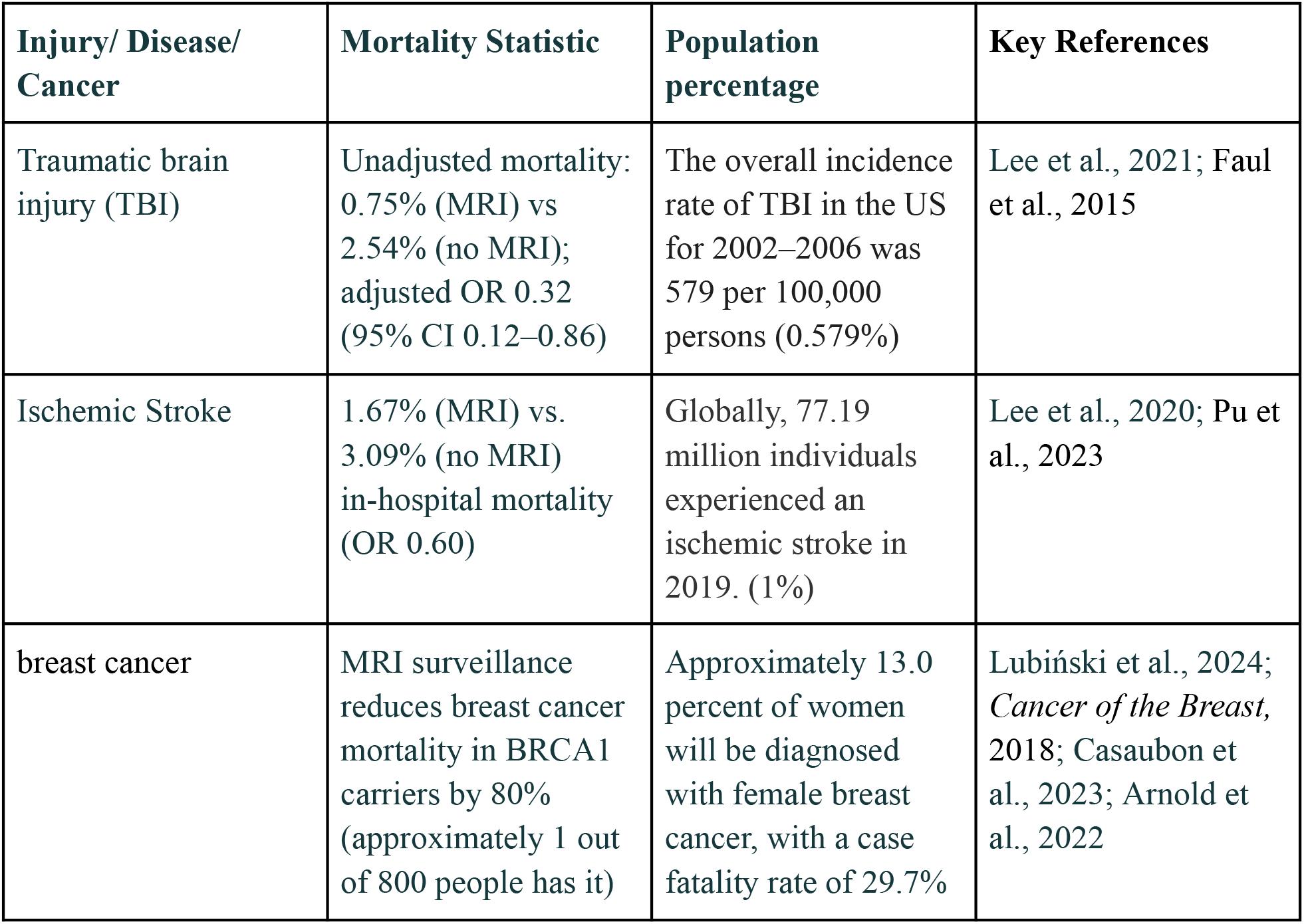
Summary of the mortality Improvement through clinical implications of MRI: focusing on breast cancer, traumatic brain injury (TBI), and Ischemic Stroke.

According to Aquino, *α* (Aquino & Ramírez-Rondán, 2019), the output elasticity of capital is estimated in the range of 0.46 and 0.56. In our research, we will substitute *α* = 0.46, 0.56, and 0.51 (the average of the maximum value and the minimum value) into the production function and compare the estimation results. The overall level of technology and efficiency in the production process, A, and the value of physical assets used in production, K, will be canceled out over the calculation process, so the real-world value of the two variables will be insignificant in this research.

### 3.3 Parameter/Variable Estimation

To estimate the aggregate mortality rate of the three conditions, given the assumption and data on 3.2, we calculated the aggregate of the incidence rate multiplied by the case-fatality rate:

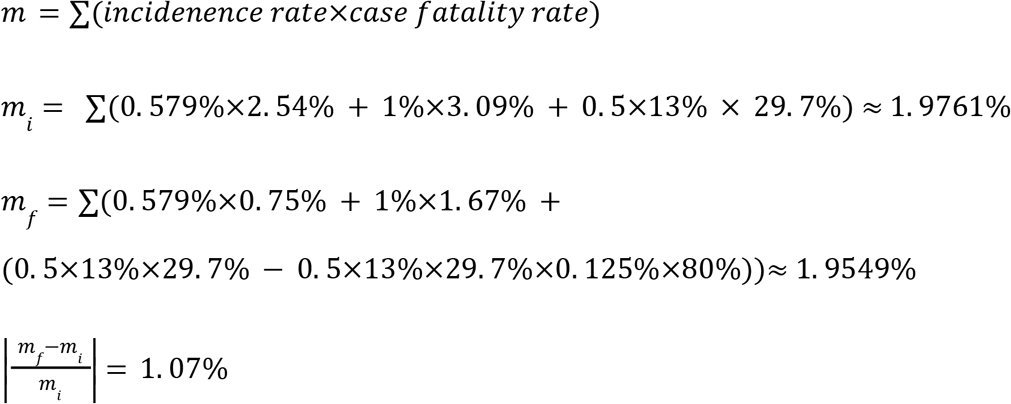

Given that *L* (*m*)= *E* (1−*m*), the labor composite *L*_*i*_ = *L* (*m*_*i*_) and *L*_*f*_=*L* (*m*_*f*_) will be:

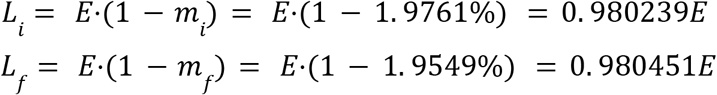

Substitute *L*_*i*_ and *L*_*f*_ back into the Cobb-Douglas production function. Given *α* = 0.46, 0.56, and 0.51, the Cobb-Douglas production function is equivalent to:

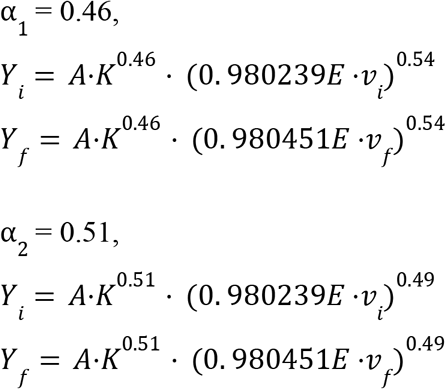

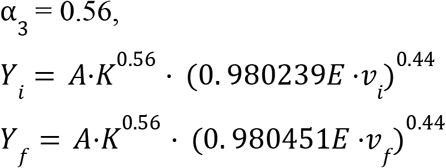

Given the calculation assumption that *v*_*f*_ = *v*_*i*_, we can do a conservative estimation on the relative percentage change of GDP after the implementation of MRI in a hypothetical developing country:

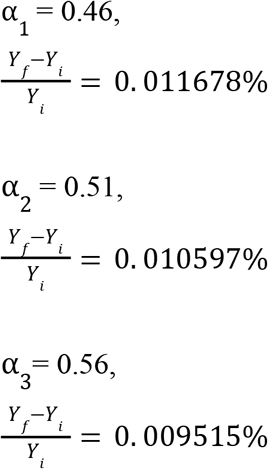

## 4. Results

### 4.1 Health Outcomes Correlation: Mortality Rate

The analysis shows that a positive correlation does exist and is significant between increased access to magnetic resonance imaging (MRI) and health outcomes at the nationwide level. The greater access to MRI, as outlined in Section 2.2, will enhance diagnostic accuracy, earlier disease identification, and reduce health disparity; all of which will serve to improve the health of the population. The availability of MRI, based on the conservative estimate that follows, can save the national mortality rate by **1.07 percent**. This quantitative result supports the qualitative evidence that has been obtained from the previous research, that MRI not only strengthens the diagnostic power of the healthcare system, but also reflects a tangible improvement in the survival rates. Basically, MRI has a direct impact on reducing preventable mortality, and, as such, the general health condition of the country is improved. These findings also support the already existing agreement in previous studies that high-quality MRI accessibility is significantly linked to positive health outcomes.

### 4.2 MRI Accessibility and GDP Calculations

According to conservative estimation under strict assumptions, the gross domestic product incremental gains due to increased MRI accessibility range from **0.009515%** to **0.011678%**, depending on the parameter of capital elasticity assumed in the Cobb–Douglas production function. Although the estimated growth is, in macroeconomic terms, a minor or even negligible improvement to the economy, it must be viewed in the context of highly restrictive assumptions. Only three clinical conditions have been included in the analysis: breast cancer, traumatic brain injury, and ischemic stroke. Considering miscellaneous diseases and health conditions, the actual mortality rate improved by increased MRI access is expected to be higher, thus implying that the GDP growth statistics are a lower-bound measure.

## 5. Discussion

### 5.1 Economic Interpretation

The conservative estimate given in this paper supports the positive correlation between the accessibility of MRI and the national economic output. Although the direct short-term payoff (within the time frame of 12 months to 2 years) may be considered comparatively small, at about **0.011%** of GDP growth, there is evidence that healthcare gains, such as MRI access, play a role in economic performance. Expanding the application to include more diseases, injuries, and health-related issues, and assuming that the post-MRI, health-adjusted labor ratio (*V*_*f*_) is above its original (*V*_*i*_), the percentage GDP growth is expected to be higher than the estimation in the research, yet would not yield exponential short-term returns.

According to the observational study from Aslan et al., the maximum GDP growth contributed by increasing **1%** of healthcare expenditure (HCE) is estimated to be **0.499%**. The study also illustrates the bi-directional causality of HCE and GDP growth for all G7 countries. The conservative estimation of GDP growth contributed by enhanced MRI accessibility from our paper falls in the range of **0.009515%** to **0.011678%**, which is considered reasonable based on the previous study from Aslan et al.. That said, even if taking miscellaneous diseases and health conditions into account, the GDP growth contributed by MRI innovation will likely not exceed the value estimated from previous studies (0.499%), which still can’t be considered statistically significant.

However, the long-run view that is above five years is not reflected in the current model. Measures of health like Disability-Adjusted Life Years (DALY) and Quality-Adjusted Life Years (QALY), which MRI has been proven to increase, offer good reasons to expect greater long-term improvements in healthcare outcomes and thus GDP. The key point is that the chain of health and financial performances is reciprocal: better economic performance contributes to health, and better health leads to an increase in productivity and economic growth in the long run. This forms a favorable feedback between the accessibility of MRI and GDP growth. The temporary effect of MRI-induced health gains on GDP may not be very significant in the short term (within a 12-month to 2-year perspective), but in the long term, the effect is expected to be much larger. Specifically, the benefits of structure due to a stronger workforce, lower morbidity, and potentially increased productivity are expected to be accrued over time.

### 5.2 Diminishing & Constant Economic Return to Scale

It has been supported with empirical data that there is a positive correlation between the accessibility of magnetic resonance imaging (MRI) and macro-economic performance. However, the effects of MRI on aggregate output, when analyzed only in the context of health-gaining benefits, indicate a tendency of decreasing marginal returns on a Cobb-Douglas production model. The magnitude of the contribution made by a single variable to output is formally summarized by the partial derivative of Y with that variable; increasing the health-related human capital (*v*) or labor (*L*) individually will increase the output, but with decreasing marginal propensities. (e.g, if the government purchases MRIs that exceed the demand of the market, then it generates economic loss. That said, none of the countries has currently experienced this circumstance.)

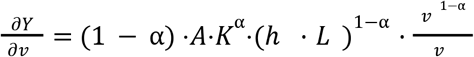

That said, the improvements in MRI technology are inseparably connected with the general technological advancements and investment. The technological breakthrough raises the total-factor productivity parameter (*A*), and the corresponding rise in the manufacturing and distribution of the MRI units adds to the capital accumulation (*K*). Once these extra channels are included, MRI-induced health gains, combined with technological and infrastructural change, could maintain a more balanced, stable return to scale on the national level.

### 5.3 Potential Policy Implications

The results are essential in relation to the policy of healthcare and the population. The empirical evidence shows that the accessibility of healthcare is positively correlated with the performance of macroeconomic programs, despite the economic return being small in the short run; therefore, governments, especially those whose economies are labor-intensive, should recognize the importance of investing in the healthcare infrastructure, such as the magnetic resonance imaging (MRI) centers. Greater access to healthcare not only reduces preventable mortality but also enhances workforce productivity in the long run, which brings about economic growth. Besides direct implementation, the state actors ought to channel resources into

MRI-related research and development. Investing in innovations like portable MRI machines, diagnostics with the help of artificial intelligence, and affordable imaging technologies will help to both make health care more accessible and guarantee a sustainable payback after some years, so that the development of both health care and the economy will work in parallel.

### 5.4 Limitations and Future Research

This research faces several important limitations. First, the data sources for MRI’s impact on mortality were restricted to only three health conditions—breast cancer, traumatic brain injury, and ischemic stroke. This narrow focus likely underestimates the full extent of MRI’s contribution to population health and, consequently, to economic output. Second, broader measures of health, such as DALY and QALY, which could be utilized to estimate the value of the variable *v*, could not be included due to data constraints, further limiting the comprehensiveness of the estimation. Third, the model assumes static conditions for several variables through the estimation process, such as employment levels, the ratio of human labor in the form of health *v* (which should be one of the independent variables in the calculation but is assumed constant due to source limitations), and comorbidity, which oversimplifies real-world dynamics. Finally, the time horizon considered is short-term (1–2 years), thereby overlooking the cumulative and potentially larger long-term benefits despite acknowledging them. Future research should, if possible, expand the scope of diseases analyzed, incorporate DALY and QALY metrics, and model dynamic, multi-year economic effects to better capture the systemic impact of MRI accessibility on national productivity.

## 6. Conclusion

This paper has explored how access to MRI has been associated with the health outcome, as well as the economic output demonstrated through a health-enhanced Cobb-Douglas production function. The conservative estimation shows that a minimum of **1.07 percent** fewer people die when access to MRI is more widespread, and given more pessimistic conditions, adds a GDP growth of **0.009515 percent** to **0.011678 percent**. Although the short-term economic benefits do not seem to be statistically substantial, the results complement the overall claim that healthcare and economic performance have a positive relationship. MRI is not only beneficial in terms of diagnostic and treatment results but also promotes potential workforce productivity and national output in the long run, which is latent in short-run macroeconomic analysis. Although short-term returns fade when either is taken separately, MRI-based technological advances, along with expanded capital and productivity gains, provide a long-term, sustainable direction toward economic and health gains.

## Data Availability

All data produced in the present study are available upon reasonable request to the authors.
All data produced in the present work are contained in the manuscript

